# Diagnostic Accuracy of MRI Radiomics for Predicting KRAS Mutation in Rectal Cancer: A Systematic Review and Meta-analysis

**DOI:** 10.64898/2026.07.17.26358357

**Authors:** Maab M. Saleh, Mahmoud Hegazy, Mohamed A. Alsaied, Aya J. Elkenani, Reem Ehab, Mohamed Hesham, Hassan Magdy Abdelrazek, Sedigheh Nazemi, Mostafa Shalaby, Alaa El-Hussuna

**Author notes:** **Corresponding to:** Mohammed A. Alsaied ***Affiliation:*** Faculty of Medicine, Mansoura University, Mansoura, Egypt, ***Telephone:***, ***Address:*** Elgomhouria Street, Mansoura City, Dakahlia Governorate, Egypt, postal code 35516, ***e-mail:***. These authors contributed equally to this work and share first authorship (Maab M. Saleh, Mahmoud Hegazy).

## Abstract

**Background:** KRAS mutation status is an important biomarker in rectal cancer, with implications for prognosis and treatment response. MRI-based radiomics has emerged as a non-invasive approach for predicting tumor genotypes. However, the diagnostic performance of MRI radiomics for predicting KRAS mutation status remains unclear. This study aimed to evaluate the diagnostic accuracy of MRI radiomics for predicting KRAS mutations in rectal cancer.

**Methods:** A systematic search of PubMed, Cochrane Library, Scopus, and Web of Science was performed through July 2025. Diagnostic test accuracy studies evaluating MRI-based radiomics or artificial intelligence models for predicting KRAS mutation status in adult patients with rectal cancer were included, using molecular testing as the reference standard. Risk of bias was assessed using the QUADAS-2 tool. Pooled sensitivity and specificity were estimated using a bivariate random-effects model.

**Results:** Seven studies involving 1,224 patients were included. The pooled sensitivity was 0.736 (95% CI: 0.697–0.772) and the pooled specificity was 0.645 (95% CI: 0.586–0.701). The false positive rate was 0.355 (95% CI: 0.299–0.414). The area under the hierarchical summary receiver operating characteristic curve was 0.754, with a normalized partial AUC of 0.666. Between-study heterogeneity ranged from low to moderate depending on the estimation method (I² = 8.4%–53.3%).

**Conclusion:** MRI radiomics demonstrates moderate diagnostic accuracy for predicting KRAS mutation status in rectal cancer and may serve as a promising non-invasive biomarker for preoperative molecular stratification. Further large-scale studies with external validation are required to confirm its clinical utility.

**Key points and Clinical relevance statement:** *Question:* Can MRI radiomics noninvasively predict Kirsten rat sarcoma(KRAS) viral oncogene homolog mutation status in rectal cancer before treatment planning?

*Findings:* In seven studies including 1,224 patients, MRI radiomics showed moderate accuracy for mutation prediction, with 0.74 sensitivity and 0.65 specificity.

*Clinical Relevance Statement:* Preoperative MRI radiomics offers a noninvasive complement to tissue testing for molecular stratification in rectal cancer, supporting treatment planning and reducing the risk of underrepresenting whole-tumor heterogeneity.

## Introduction

For rectal cancer, the pressure point is neither solely anatomical staging nor just biological staging. It is the merging of biological factors into preoperative decision-making. Of all the molecular changes with immediate clinical relevance, KRAS is of particular interest due to its role in determining tumor behavior, its predictive value for anti-EGFR therapy resistance in metastatic colorectal cancer, and its association with negative oncologic outcomes in neoadjuvant-treated rectal cancer cohorts [1–3]. Nonetheless, KRAS status is predominantly ascertained via tissue sampling, which offers a single, limited, and spatially heterogeneous tumor sample and is unsuitable for the repetitive, whole-tumor evaluation across the continuum of care [4, 5].

This justifies the case for imaging-based genotype surrogates. Pelvic MRI is an integral part of managing rectal cancer, as it provides information on tumor extent, involvement of the mesorectal fascia, extramural venous invasion, nodal disease, and response anatomy, which impact multimodal treatment planning [6, 7]. The next logical step is to determine whether the same examination can obtain molecular data, thereby expanding its utility from a staging test to one that integrates imaging for phenotypic and genotypic risk stratification.

Radiomics provides that opportunity. Since the pioneering radiomics framework proposed by Lambin et al. (2012) [8] and further described by Gillies et al. (2016) [9], the scientific community has focused on developing automated methods to mine previously inaccessible information from standard-of-care imaging. Radiomic signatures can, in theory, represent the imaging phenotype of the molecular program and correlate with intratumoral heterogeneity, microenvironmental structuring, and molecular imaging [10, 11]. However, interest has been tempered by concerns regarding segmentation variability, scanner dependence, and feature instability, overfitting, and inadequate external validation, precisely the issues that the Image Biomarker Standardization Initiative and subsequent methodological recommendations sought to address [12].

With regard to rectal cancer, some MRI studies offer promising, yet variable, results on the performance of texture analysis, handcrafted radiomics, and machine-learning classifiers focused on T2-weighted or multiparametric MRI for preoperative KRAS prediction, though the studies are often fragmented and methodologically diverse, and so difficult to translate into clinically meaningful diagnostic predictions [13–15]. As such, there is a need for focused synthesis around diagnostic accuracy. This study aimed to evaluate the diagnostic accuracy of MRI radiomics for predicting KRAS mutations in rectal cancer.

## Methods

We conducted this systematic review and meta-analysis in accordance with the Preferred Reporting Items for Systematic Reviews and Meta-Analyses (PRISMA) guidelines and the recommendations outlined in the Cochrane Handbook for Systematic Reviews of Diagnostic Test Accuracy [16, 17].

### Search strategy and information sources

A comprehensive literature search was performed in PubMed, the Cochrane Library, Scopus, and Web of Science from database inception through July 2025. The search strategy combined controlled vocabulary and keywords related to rectal cancer, magnetic resonance imaging, artificial intelligence, and KRAS mutations.

The following Medical Subject Headings (MeSH) terms were used: *“Rectal Neoplasms,” “Magnetic Resonance Imaging,” “Artificial Intelligence,” “Machine Learning,” “Genes, KRAS,”* and *“Proto-Oncogene Proteins p21(ras)”*, which were combined using Boolean operators to develop database-specific search queries.

Only studies published in English were included. No additional restrictions or filters were applied in order to maximize sensitivity and minimize the risk of missing relevant studies. A deliberately broad search strategy was adopted to maximize sensitivity and ensure that all potentially relevant studies were captured.

The complete search strategies for each database are provided in **Table S1** in the Supplementary Materials.

### Inclusion criteria

We incorporated studies meeting all of these criteria: ***(1) Population*** – patients aged 18 years and older with rectal cancer confirmed through histopathological evaluation; ***(2) Index test –*** application of radiomics-based artificial intelligence models on pre-treatment MRIs for predicting KRAS mutations of the lesions; ***(3) Reference standard –*** KRAS mutation status ascertained through recognized molecular diagnostic techniques including but not limited to PCR, NGS, or Sanger sequencing; and ***(4) Design –*** either prospective or retrospective studies focused on diagnostic test accuracy with at least one performed or calculable diagnostic accuracy metric (e.g. sensitivity, specificity, area under the ROC curve, or diagnostic odds ratio).

### Exclusion criteria

All study designs were accepted except reviews, letters to editors, abstracts, opinions, non-human studies, and non-English studies. Studies that used radiomics other than MRI-based radiomics, or cancers other than rectal cancer, were also excluded.

### Study Selection

After exporting search results, articles were imported into the Rayyan platform [18] for manual detection and deletion of duplicate records. A two-step screening process was undertaken: the first step involved screening titles, abstracts, and keywords, and the second step involved reviewing the full text of all retrieved studies to determine final eligibility. The articles were evaluated by two independent authors (M.H. and R.E.). Conflicts between the two authors were resolved through discussion or consultation with an independent third author (A.E.).

### Data extraction

Two authors (A.J.E. and H.M.A.) manually extracted the data and entered it into a standardized form in Google Sheets, and the data were checked by a third author (A.E.). Study characteristics, including the name of the first author and year of publication, the study design, and the imaging modality, were extracted. AI model-related information was also extracted from each study, including the classifier and feature-extraction software, the model employed, the segmentation technique, and the validation strategy.

### Risk of bias assessment

The quality of the included studies was assessed using the revised Quality instrument. Assessment of Diagnostic Accuracy Studies 2 (QUADAS-2) tool [19] by two independent authors (M.A.S and M.M.S). The Quality Assessment of Diagnostic Accuracy Studies 2 (QUADAS-2), which includes four domains: patient selection, index test, reference standard, and flow & timing, was used to evaluate the risk of bias of the individual studies. Any disagreements were resolved through discussion, and if no consensus was achieved, a third author’s (M.S) decision was sought.

### Statistical analysis, Subgroups and Heterogeneity

Diagnostic performance metrics—sensitivity, specificity, accuracy, and estimated disease prevalence—were extracted or calculated from each included study, then arranged into standard 2×2 confusion tables specifying true positives (TP), false positives (FP), true negatives (TN), and false negatives (FN) according to the principles of Monaghan et al. 2021 study [20]. These tables formed the baseline for all subsequent pooled analyses.

The primary meta-analysis employed a bivariate random-effects model, which synthesizes sensitivity and specificity simultaneously while acknowledging their within-study correlation. This framework provides pooled point estimates and 95 % confidence intervals, accommodating both sampling error and variability across studies [21].

All statistical methods were conducted on R software (version 4.4.2, 2024) [22] using “mada” and “meta” packages. A Hierarchical Summary Receiver Operating Characteristic (HSROC) curve derived from the bivariate model graphically depicts the trade-off between sensitivity and specificity across all investigations [23]. The area under the HSROC curve (AUC) and normalized partial AUC summarize the overall accuracy of the artificial-intelligence models under review. In parallel with the bivariate analysis, separate univariate random-effects models were fitted for sensitivity and specificity. Forest plots display estimates from each study alongside pooled values and their 95-percent confidence intervals, providing an alternative view of diagnostic performance.

Assessment of between-study heterogeneity used two complementary methods: the Zhou-Dendukuri model [24], which incorporates covariance between sensitivity and specificity, and the Holling-adjusted I², a bias-corrected variance estimate common in diagnostic test accuracy research. Heterogeneity was graded according to standard benchmarks, with I² around 25% regarded as low, 50% as moderate, and 75% as high.

## Results

### Study Selection

Database search identified 948 studies. The screening process ultimately identified seven studies (Meng et al., (2019) [25], Oh et al., (2020) [26], Cui et al., (2020) [14], Guo et al., (2020) [27], Zhang ZY et al., (2021) [15], Zhang G et al., (2021) [28], and Xiang et al., (2023) [29]) meeting the eligibility criteria, which were incorporated into the systematic review and quantitative pooled analysis. Thes studies were conducted between 2019 and 2023. A flow diagram illustrating the results of the study selection process is shown in Figure 1.

**Figure 1.**
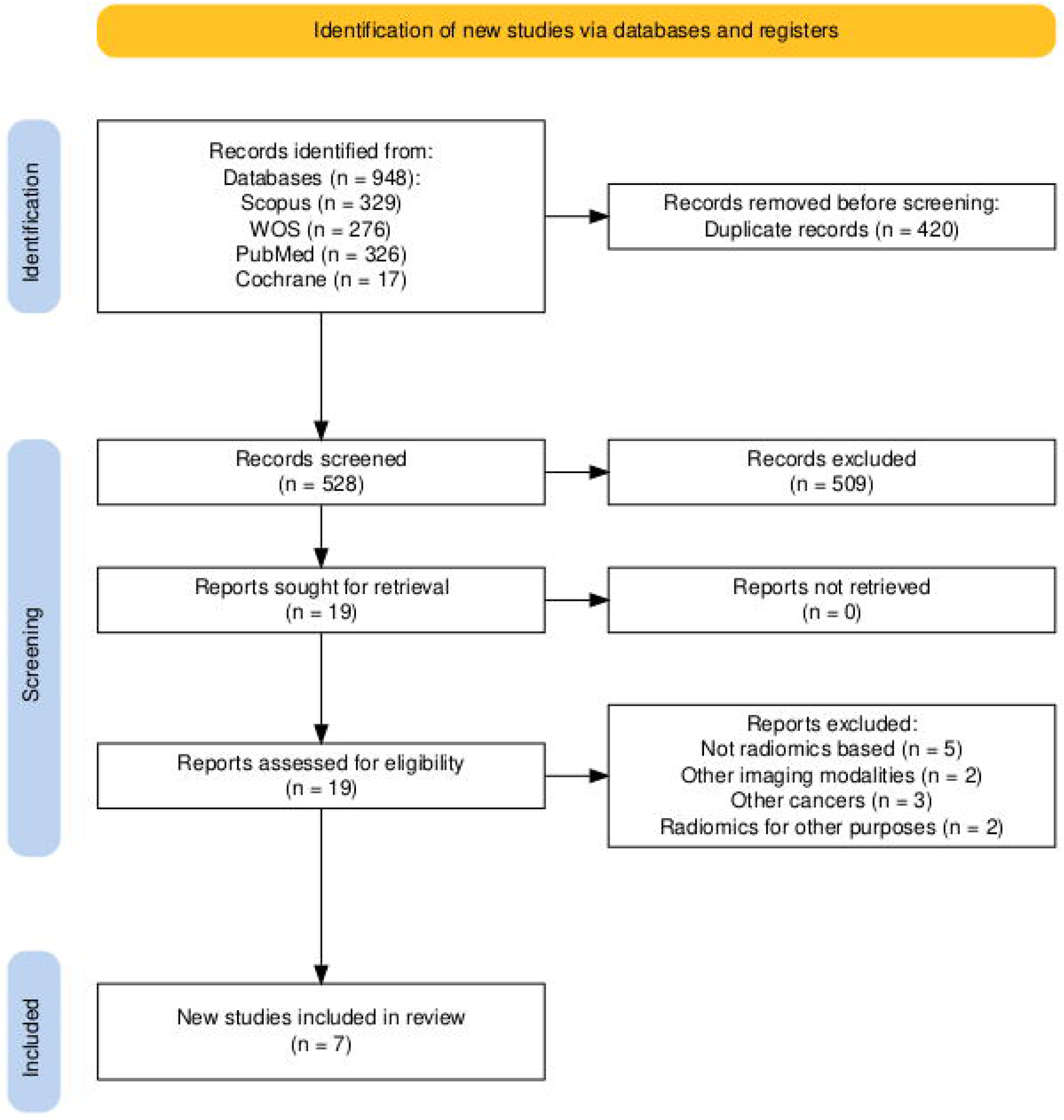
PRISMA 2020 flow diagram of study selection.

### Characteristics of included studies

The seven studies used various modalities of magnetic resonance imaging (MRI); some studies used T2-weighted imaging (T2WI) [14, 15, 29], and the remaining used T2WI + Diffusion weighted imaging (DWI) [26–28] or multiparametric MRI [25]. Feature selection using LASSO, MRMR, PCA, ANOVA/KW, or recursive feature elimination was used. Artificial intelligence models used also differed between studies, with some using support vector machines (SVMs) [14, 25, 26, 29], random forests [25, 26], LASSO regression [15, 25, 29], and advanced deep learning models such as 3D V-Net or ensemble models [28]. Most segmentations were manual, although one study compared deep-learning segmentation with manual segmentation.

Preprocessing steps often included normalization, standardization, independent cohorts, internal-external validation, or predefined train/test splits. Table 1 shows a detailed summary of the included studies.

**Table 1.** Summary of included studies.

| Study | Aim | Population | Imaging Modalities | AI Model Used | Segmentation Method | Preprocessing Steps | Feature Selection Method | Validation Method |
| --- | --- | --- | --- | --- | --- | --- | --- | --- |
| **Meng et al. 2019** | Predict biological characteristics (LN metastasis, HER2, Ki-67, KRAS) using MP-MRI | patients who underwent radical resection for rectal cancer | Multiparametric MRI | Random Forest, SVM, LASSO | Manual | Normalization and reproducibility check | MRMR + Wilcoxon | Independent validation |
| **Oh et al. 2020** | Differentiate between tumors with mutant KRAS and wild-type KRAS | collected from the tumor bank or prospectively from patients with mid to lower rectal cancer | MRI (T2WI + DWI) | Ensemble (Random Forest + SVM) | Manual | Standardization and normalization | PCA | Train/Test split (70/30) |
| **Cui et al. 2020** | Predict KRAS mutation status | Pathological confirmed Rectal cancer diagnosis | MRI (T2WI) | SVM | Manual | Normalization, dynamic range limiting | ANOVA/KW + Logistic Regression | Internal + External validation |
| **Guo et al. 2020** | Predict KRAS mutation status | Diagnosed with rectal cancer receiving no prior cancer treatment with available KRAS status | MRI (T2WI + DWI) | Random Forest | Manual | Normalization | Recursive Feature Elimination | Train/Test split |
| **Zhang ZY et al. 2021** | Predict KRAS status | Histologically confirmed rectal adenocarcinoma | MRI (T2WI) | LASSO regression | Manual | None detailed | LASSO | 7:3 split (train/test) |
| **Zhang G et al. 2021** | Predict KRAS/NRAS/BR AF mutations | Histologically confirmed rectal adenocarcinoma | MRI (T2WI, DWI and T2WI+DWI) | Deep Learning (3D V-Net) | DL-based and manual segmentation compared | None detailed | Not clear, radiomics-based models | Train/Validation split |
| **Xiang et al. 2023** | Predict KRAS mutation status | Histologically confirmed rectal adenocarcinoma with available KRAS status | MRI (T2WI) | SVM + LASSO | Manual | Radiomics extraction | LASSO | Train/test split |

MRI: Magnetic resonance imaging, SVM: Support Vector Machine, LASSO: Least Absolute Shrinkage and Selection Operator, DWI: Diffusion-Weighted Imaging, DL: Deep Learning

### Risk of bias assessment

Quality assessment of included studies was done using QUADAS -2 tool. Assessment revealed that five studies (Xiang et al. 2023 [29], Oh et al. 2020 [26], Guo et al. 2023 [27], Zhang ZY et al. 2021 [15] and Zhang G et al. 2021 [28]) showed low risk of bias. Moreover, the remaining two studies (Meng et al. 2019 [25] and Cui et al. 2020 [14]) demonstrated some concerns. Figure 2 shows risk of bias graph of included studies.

**Figure 2.**
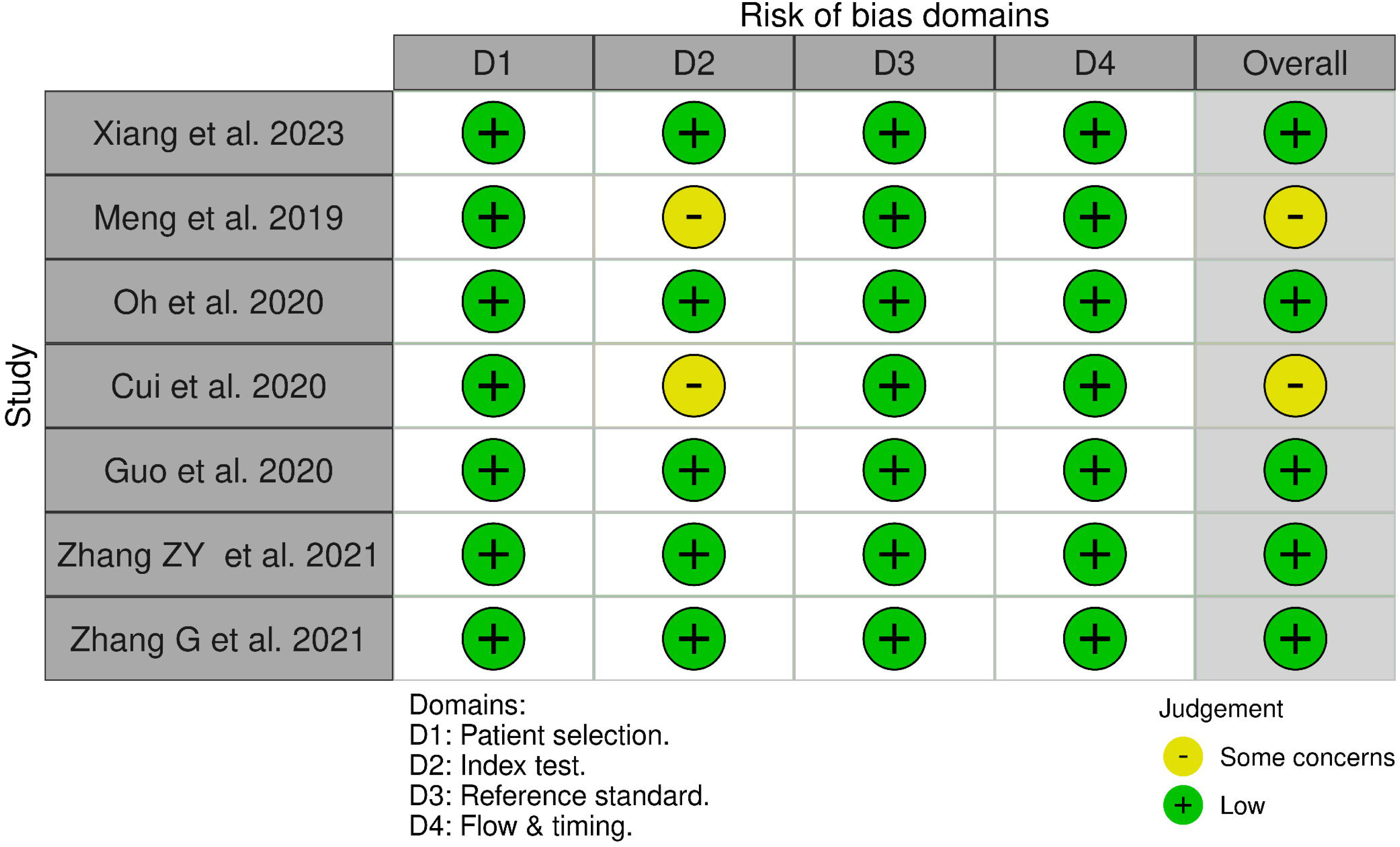
QUADAS-2 risk of bias and applicability assessment. Risk of bias was evaluated across four domains: D1, patient selection; D2, index test; D3, reference standard; and D4, flow and timing.

### Diagnostic test accuracy

A diagnostic-test-accuracy meta-analysis was conducted to assess how well artificial-intelligence radiomics models predict KRAS mutation in rectal cancer from MRI data. Across all cohorts, the pooled sensitivity was 0.736 (95% CI = [0.697, 0.772]) and specificity was 0.645 (95% CI = [0.586, 0.701]), corresponding to a false positive rate (FPR) of 0.355 (95% CI = [0.299, 0.414]). The overall area under the curve (AUC) was 0.754, with a normalized partial AUC (restricted to the range of observed FPRs) of 0.666. The hierarchical summary receiver operating characteristic (HSROC) curve for all cohorts is presented in Figure 3, and forest plots of the univariate analyses of sensitivity and specificity are shown in Figure 4. Between-study heterogeneity was assessed using the Zhou and Dendukuri approach, revealing low heterogeneity (I² = 8.4%), while the Holling adjusted method estimated heterogeneity in the range of 2.0% to 2.2%.

**Figure 3.**
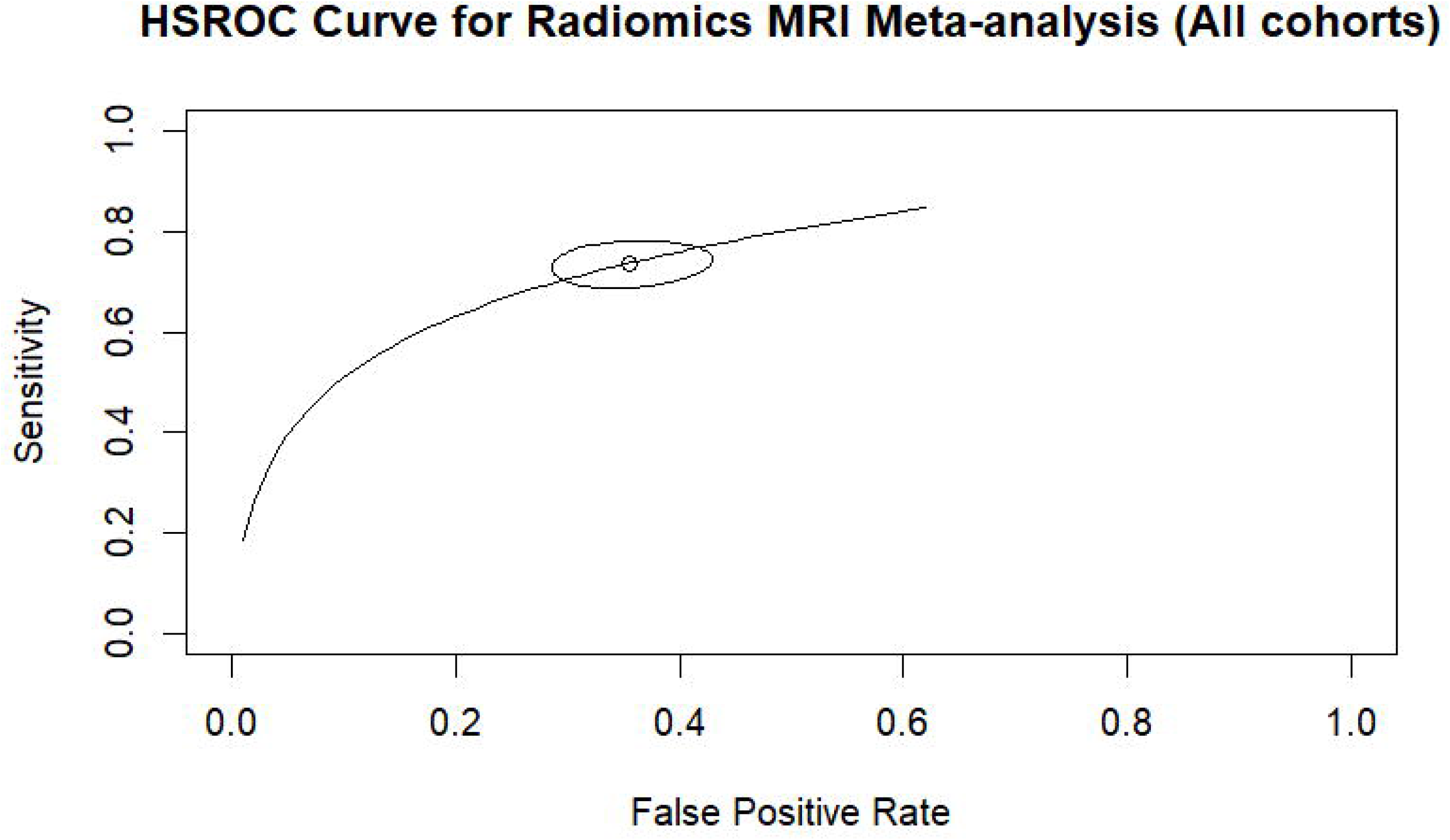
HSROC curve of AI-based MRI radiomics for predicting KRAS mutation in rectal cancer (all cohorts). The open circle denotes the summary operating point of the meta-analysis, while the surrounding ellipse indicates the uncertainty around the pooled estimate.

**Figure 4.**
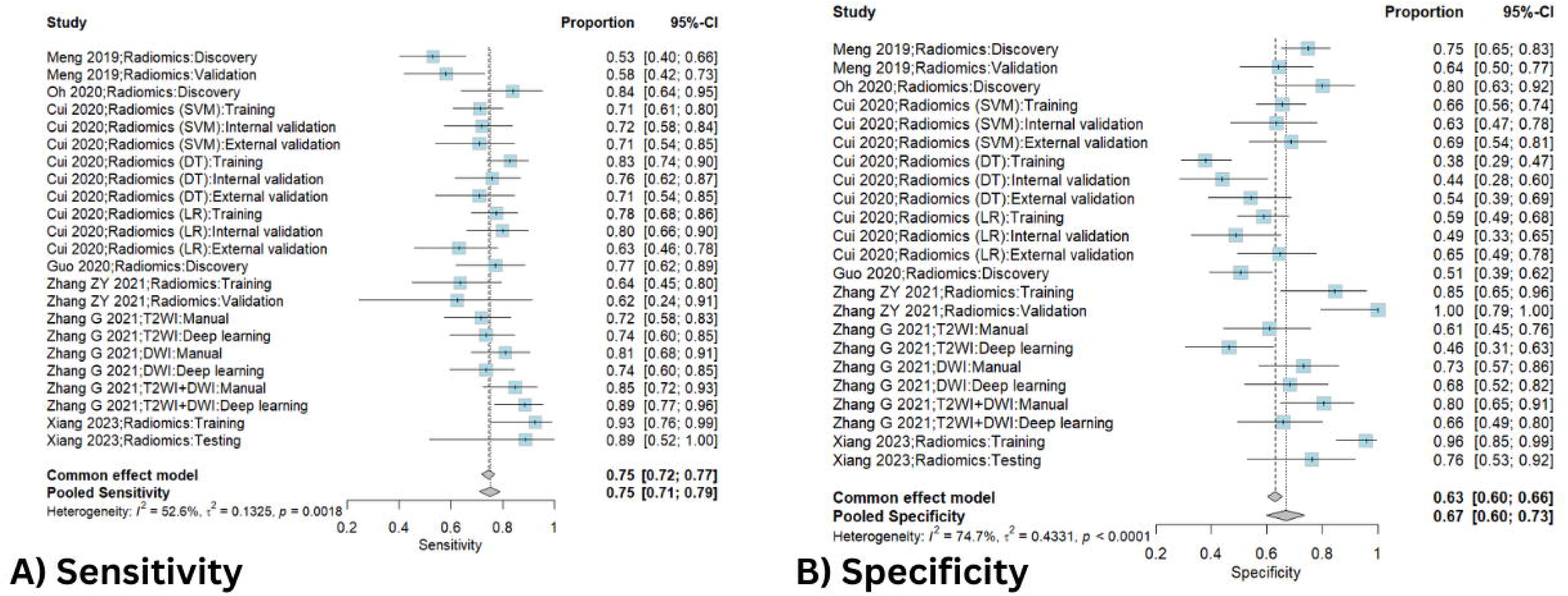
Forest plots of sensitivity and specificity for AI-based MRI radiomics predicting KRAS mutation (all cohorts). Panel A shows study-level and pooled estimates of sensitivity, and Panel B shows study-level and pooled estimates of specificity.

When restricting the analysis to validation cohorts only, the pooled sensitivity remained stable at 0.737 (95% CI = [0.680, 0.787]) and specificity slightly increased to 0.653 (95% CI = [0.597, 0.705]), with a corresponding FPR of 0.347 (95% CI = [0.295, 0.403]). The pooled AUC for validation datasets was 0.744, and the normalized partial AUC was 0.566. The HSROC curve for the validation cohorts is shown in **Figure 5**, and univariate forest plots for sensitivity and specificity are shown in **Figure 6**. Heterogeneity remained low in this subgroup, with I² = 5.5% based on the Zhou and Dendukuri method and ranged from 2.9% to 3.1% using the Holling adjustment method.

**Figure 5.**
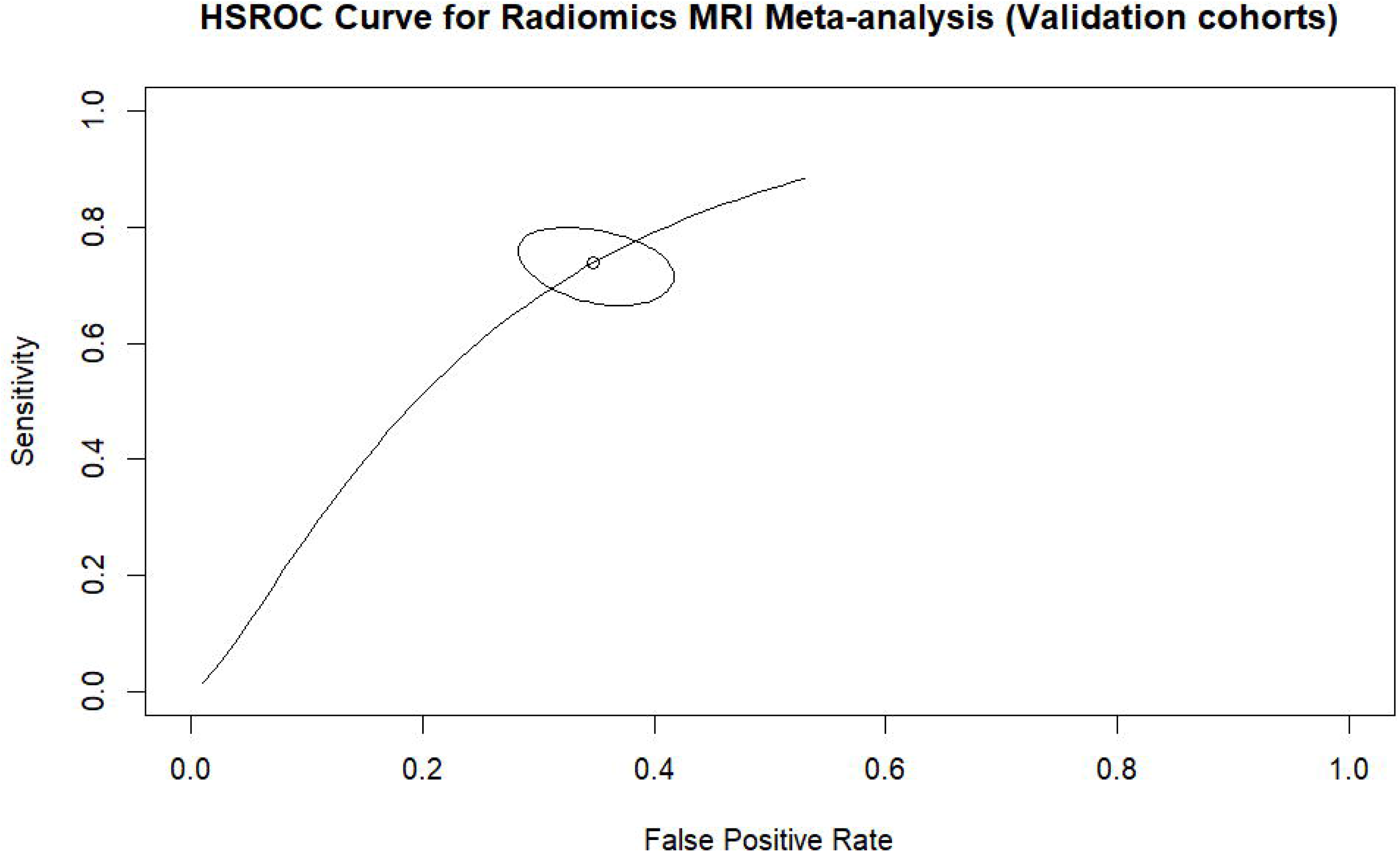
HSROC curve of AI-based MRI radiomics for predicting KRAS mutation (validation cohorts). The open circle denotes the summary operating point of the meta-analysis, while the surrounding ellipse indicates the uncertainty around the pooled estimate

**Figure 6.**
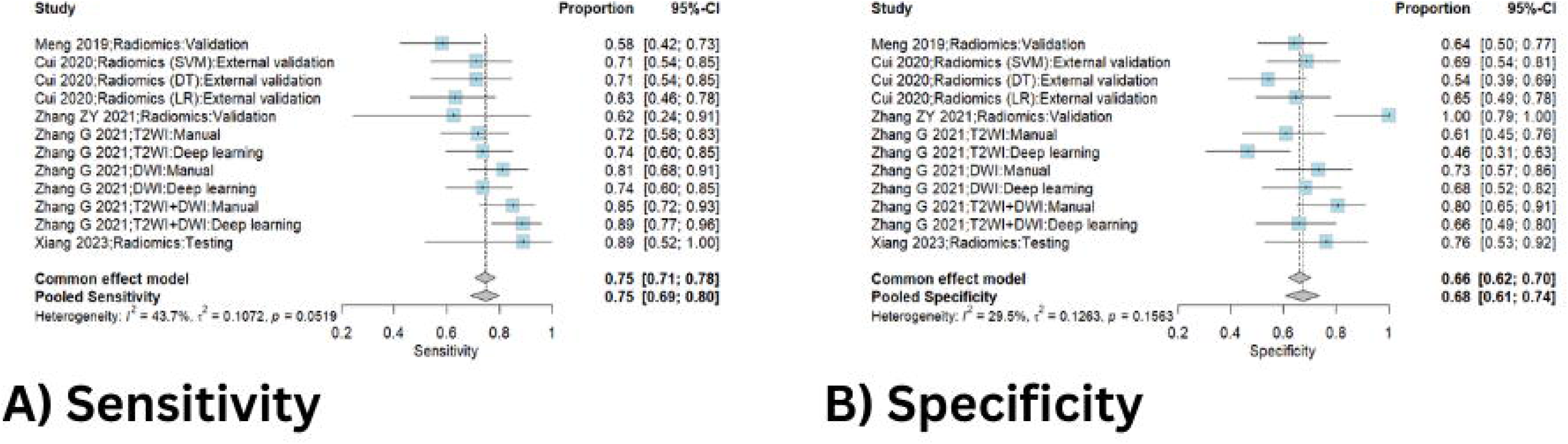
Forest plots of sensitivity and specificity for AI-based MRI radiomics predicting KRAS mutation (validation cohorts). Panel A shows study-level and pooled estimates of sensitivity, and Panel B shows study-level and pooled estimates of specificity.

## Discussion

The present systematic review and bivariate diagnostic accuracy meta-analysis evaluated the performance of MRI-based radiomics models for predicting KRAS mutation status in rectal cancer.

Across seven studies involving 1,224 patients, MRI radiomics demonstrated moderate diagnostic accuracy, with pooled sensitivity of 0.736, specificity of 0.645, and an HSROC AUC of 0.754.

These findings indicate that MRI-derived radiomic features contain measurable imaging signatures associated with KRAS mutation status, supporting the emerging concept of radiogenomics, in which imaging phenotypes reflect underlying molecular alterations.

The moderate yet consistent diagnostic performance observed across studies suggests that MRI radiomics may serve as a non-invasive adjunct biomarker for preoperative molecular characterization in rectal cancer. Given that KRAS status is currently determined through tissue-based molecular testing, which may suffer from sampling bias and cannot easily be repeated, imaging-based prediction methods could provide a complementary tool for whole-tumor assessment.

Our results are comparable to broader systematic evaluations of artificial intelligence models for KRAS prediction in colorectal cancer. A recent meta-analysis reported pooled sensitivity of 0.78 and specificity of 0.73 for MRI-based models predicting KRAS mutations, further supporting the moderate diagnostic performance of imaging-based radiogenomic approaches [30].

Several factors may explain the moderate diagnostic accuracy observed across studies.

First, methodological heterogeneity among radiomics pipelines likely contributes to variability in model performance. The included studies used diverse feature extraction methods, segmentation strategies, and machine learning algorithms, including support vector machines, logistic regression, and random forest classifiers. Such variability may influence the reproducibility of radiomic signatures across datasets.

Second, MRI protocol differences may affect feature stability. Some studies relied solely on T2-weighted sequences, while others incorporated multiparametric MRI including diffusion-weighted imaging. Previous research has shown that combining multiple MRI sequences can improve the predictive capability of radiomics models by capturing complementary biological information.

Third, limited external validation remains a common issue in radiomics research. Many studies rely primarily on internal validation cohorts, which may lead to optimistic estimates of diagnostic performance. External validation across multiple institutions is essential to ensure generalizability and clinical applicability.

Finally, the biological relationship between imaging phenotypes and specific genetic mutations such as KRAS is inherently complex. Radiomic features may capture tumor heterogeneity and microenvironmental characteristics indirectly associated with genetic alterations, but these imaging signatures may not fully represent the underlying molecular mechanisms.

Despite these limitations, MRI radiomics offers several potential clinical advantages. Unlike tissue-based molecular testing, imaging analysis is non-invasive, repeatable, and capable of evaluating the entire tumor volume, potentially reducing the risk of sampling bias associated with biopsy. Radiomics-based models could therefore complement molecular diagnostics by providing preoperative insights into tumor biology.

If further validated, MRI radiomics may contribute to preoperative risk stratification, treatment planning, and individualized management strategies for rectal cancer patients.

Several limitations should be acknowledged. First, the number of included studies was relatively small, reflecting the emerging nature of radiomics research in KRAS prediction. Second, most studies were retrospective and single-center, which may introduce selection bias and limit generalizability. Third, heterogeneity in imaging protocols, feature extraction methods, and machine learning algorithms may have influenced pooled estimates of diagnostic accuracy.

Additionally, many radiomics models lack standardized reporting and external validation, which remains a major barrier to clinical translation. Future studies should prioritize multicenter datasets, standardized radiomics workflows, and transparent reporting frameworks such as the Image Biomarker Standardization Initiative (IBSI) [12].

Future research should focus on improving the robustness and clinical applicability of radiomics models. Integrating radiomics features with clinical variables, genomic data, and deep learning approaches may enhance predictive performance. Furthermore, prospective multicenter studies with standardized imaging protocols and independent validation cohorts are needed to confirm the reproducibility of radiomics-based KRAS prediction.

Advances in automated segmentation, harmonization of imaging protocols, and radiomics standardization may further facilitate the translation of radiogenomic models into clinical practice.

## Conclusion

MRI-based radiomics demonstrates moderate diagnostic accuracy for predicting KRAS mutation status in rectal cancer. The pooled results of this diagnostic test accuracy meta-analysis suggest that radiomic features extracted from preoperative MRI contain imaging signatures associated with KRAS mutational status, supporting the concept of radiogenomic correlation in rectal cancer. As a non-invasive and repeatable imaging approach, MRI radiomics may provide complementary information for preoperative molecular stratification and treatment planning.

However, the current evidence remains limited by methodological heterogeneity and the predominance of retrospective single-center studies. Further large-scale, multicenter investigations with standardized radiomics workflows and external validation are required to confirm the robustness and clinical utility of MRI radiomics for KRAS mutation prediction.

## Data Availability

All data produced in the present study are available upon reasonable request to the authors

## Abbreviations list

AI: artificial intelligence
ANOVA/KW: analysis of variance / Kruskal-Wallis
AUC: area under the curve
CI: confidence interval
D1: patient selection
D2: index test
D3: reference standard
D4: flow and timing
DWI: diffusion-weighted imaging
EGFR: epidermal growth factor receptor
FN: false negative
FP: false positive
FPR: false positive rate
HSROC: hierarchical summary receiver operating characteristic
I²: heterogeneity statistic
IBSI: Image Biomarker Standardization Initiative
KRAS: Kirsten rat sarcoma viral oncogene homolog
LASSO: least absolute shrinkage and selection operator
MeSH: Medical Subject Headings
MRI: magnetic resonance imaging
MRMR: minimum redundancy maximum relevance
NGS: next-generation sequencing
PCA: principal component analysis
PCR: polymerase chain reaction
PRISMA: Preferred Reporting Items for Systematic Reviews and Meta-Analyses
QUADAS-2: Quality Assessment of Diagnostic Accuracy Studies 2
ROC: receiver operating characteristic
SVM: support vector machine
T2WI: T2-weighted imaging
TN: true negative
TP: true positive

## References

1. Peng J, Lv J, Peng J (2021) KRAS mutation is predictive for poor prognosis in rectal cancer patients with neoadjuvant chemoradiotherapy: a systemic review and meta-analysis. Int J Colorectal Dis 36:1781–1790. 10.1007/s00384-021-03911-z

2. Zhu G, Pei L, Xia H, et al (2021) Role of oncogenic KRAS in the prognosis, diagnosis and treatment of colorectal cancer. Mol Cancer 20:143. 10.1186/s12943-021-01441-4

3. Takeda M, Yoshida S, Inoue T, et al (2025) The Role of KRAS Mutations in Colorectal Cancer: Biological Insights, Clinical Implications, and Future Therapeutic Perspectives. Cancers 17:428. 10.3390/cancers17030428

4. Petaccia de Macedo M, Melo FM, Ribeiro HSC, et al (2017) KRAS mutation status is highly homogeneous between areas of the primary tumor and the corresponding metastasis of colorectal adenocarcinomas: one less problem in patient care. Am J Cancer Res 7:1978–1989

5. Cefalì M, Epistolio S, Palmarocchi MC, et al (2021) Research progress on *KRAS* mutations in colorectal cancer. J Cancer Metastasis Treat 7:N/A-N/A. 10.20517/2394-4722.2021.61

6. Beets-Tan RGH, Lambregts DMJ, Maas M, et al (2018) Magnetic resonance imaging for clinical management of rectal cancer: Updated recommendations from the 2016 European Society of Gastrointestinal and Abdominal Radiology (ESGAR) consensus meeting. Eur Radiol 28:1465–1475. 10.1007/s00330-017-5026-2

7. Lee S, Kassam Z, Baheti AD, et al (2023) Rectal cancer lexicon 2023 revised and updated consensus statement from the Society of Abdominal Radiology Colorectal and Anal Cancer Disease-Focused Panel. Abdom Radiol 48:2792–2806. 10.1007/s00261-023-03893-2

8. Lambin P, Rios-Velazquez E, Leijenaar R, et al (2012) Radiomics: extracting more information from medical images using advanced feature analysis. Eur J Cancer 48:441–446. 10.1016/j.ejca.2011.11.036

9. Gillies RJ, Kinahan PE, Hricak H (2016) Radiomics: Images Are More than Pictures, They Are Data. Radiology 278:563–577. 10.1148/radiol.2015151169

10. Badic B, Tixier F, Cheze Le Rest C, et al (2021) Radiogenomics in Colorectal Cancer. Cancers 13:973. 10.3390/cancers13050973

11. Li S, Zhou B (2022) A review of radiomics and genomics applications in cancers: the way towards precision medicine. Radiat Oncol 17:217. 10.1186/s13014-022-02192-2

12. Zwanenburg A, Vallières M, Abdalah MA, et al (2020) The Image Biomarker Standardization Initiative: Standardized Quantitative Radiomics for High-Throughput Image-based Phenotyping. Radiology 295:328–338. 10.1148/radiol.2020191145

13. Shin YR, Kim KA, Im S, et al (2016) Prediction of KRAS Mutation in Rectal Cancer Using MRI. Anticancer Res 36:4799–4804. 10.21873/anticanres.11039

14. Cui Y, Liu H, Ren J, et al (2020) Development and validation of a MRI-based radiomics signature for prediction of KRAS mutation in rectal cancer. Eur Radiol 30:1948–1958. 10.1007/s00330-019-06572-3

15. Zhang Z, Shen L, Wang Y, et al (2021) MRI Radiomics Signature as a Potential Biomarker for Predicting KRAS Status in Locally Advanced Rectal Cancer Patients. Front Oncol 11:614052. 10.3389/fonc.2021.614052

16. McInnes MDF, Moher D, Thombs BD, et al (2018) Preferred Reporting Items for a Systematic Review and Meta-analysis of Diagnostic Test Accuracy Studies: The PRISMA-DTA Statement. JAMA 319:388–396. 10.1001/jama.2017.19163

17. Jonathan J. Deeks, Patrick M. Bossuyt, Mariska M. Leeflang, Yemisi Takwoingi (2023) Cochrane Handbook for Systematic Reviews of Diagnostic Test Accuracy. John Wiley & Sons, Chichester (UK)

18. Ouzzani M, Hammady H, Fedorowicz Z, Elmagarmid A (2016) Rayyan—a web and mobile app for systematic reviews. Syst Rev 5:210. 10.1186/s13643-016-0384-4

19. Whiting PF, Rutjes AWS, Westwood ME, et al (2011) QUADAS-2: a revised tool for the quality assessment of diagnostic accuracy studies. Ann Intern Med 155:529–536. 10.7326/0003-4819-155-8-201110180-00009

20. Monaghan TF, Rahman SN, Agudelo CW, et al (2021) Foundational Statistical Principles in Medical Research: Sensitivity, Specificity, Positive Predictive Value, and Negative Predictive Value. Medicina (Mex) 57:503. 10.3390/medicina57050503

21. Negeri ZF (2025) A Bivariate Finite Mixture Random Effects Model for Identifying and Accommodating Outliers in Diagnostic Test Accuracy Meta-Analyses. Biom J Biom Z 67:e70062. 10.1002/bimj.70062

22. R Core Team (2021) R: A Language and Environment for Statistical Computing

23. Rutter CM, Gatsonis CA (2001) A hierarchical regression approach to meta-analysis of diagnostic test accuracy evaluations. Stat Med 20:2865–2884. 10.1002/sim.942

24. Zhou Y, Dendukuri N (2014) Statistics for quantifying heterogeneity in univariate and bivariate meta-analyses of binary data: the case of meta-analyses of diagnostic accuracy. Stat Med 33:2701–2717. 10.1002/sim.6115

25. Meng X, Xia W, Xie P, et al (2019) Preoperative radiomic signature based on multiparametric magnetic resonance imaging for noninvasive evaluation of biological characteristics in rectal cancer. Eur Radiol 29:3200–3209. 10.1007/s00330-018-5763-x

26. Oh JE, Kim MJ, Lee J, et al (2020) Magnetic Resonance-Based Texture Analysis Differentiating KRAS Mutation Status in Rectal Cancer. Cancer Res Treat 52:51–59. 10.4143/crt.2019.050

27. Guo X, Yang W, Yang Q, et al (2020) Feasibility of MRI Radiomics for Predicting KRAS Mutation in Rectal Cancer. Curr Med Sci 40:1156–1160. 10.1007/s11596-020-2298-6

28. Zhang G, Chen L, Liu A, et al (2021) Comparable Performance of Deep Learning–Based to Manual-Based Tumor Segmentation in KRAS/NRAS/BRAF Mutation Prediction With MR-Based Radiomics in Rectal Cancer. Front Oncol 11:696706. 10.3389/fonc.2021.696706

29. Xiang Y, Li S, Song M, et al (2023) KRAS status predicted by pretreatment MRI radiomics was associated with lung metastasis in locally advanced rectal cancer patients. BMC Med Imaging 23:210. 10.1186/s12880-023-01173-5

30. Chen K, Qu Y, Han Y, et al (2025) Performance of Machine Learning in Diagnosing KRAS (Kirsten Rat Sarcoma) Mutations in Colorectal Cancer: Systematic Review and Meta-Analysis. J Med Internet Res 27:e73528. 10.2196/73528

