## Supplementary materials for "Diagnostic Accuracy of MRI Radiomics for Predicting KRAS Mutation in Rectal Cancer: A Systematic Review and Meta-analysis"

**Table S1.** Detailed database-specific search strategy

| **Database** | **Search strategy** | **Search results** |
| --- | --- | --- |
| Scopus | ALL("rectal cancer" OR "rectal carcinoma" OR "rectal neoplasm*")  AND ALL("MRI" OR "MR imaging" OR "magnetic resonance")  AND ALL(radiomic* OR "texture analysis" OR "feature extraction") | 329 |
| Web Of Science | ALL=("rectal cancer" OR "rectal carcinoma" OR "rectal neoplasm*")  AND ALL=("MRI" OR "MR imaging" OR "magnetic resonance")  AND ALL=(radiomic* OR "texture analysis" OR "feature extraction") | 276 |
| PubMed | ("Rectal Neoplasms"[Mesh] OR "rectal cancer"[All Fields] OR "rectal carcinoma"[All Fields] OR "rectal neoplasm*"[All Fields]) AND ("Magnetic Resonance Imaging"[Mesh] OR MRI[All Fields] OR "MR imaging"[All Fields] OR "magnetic resonance"[All Fields]) AND (radiomic*[All Fields] OR "texture analysis"[All Fields] OR "feature extraction"[All Fields] OR "Radiomics"[Mesh]) | 326 |
| Cochrane | ("rectal cancer" OR "rectal carcinoma" OR "rectal neoplasm*")  AND ("MRI" OR "MR imaging" OR "magnetic resonance")  AND (radiomic* OR "texture analysis" OR "feature extraction") | 17 |
